# Structural Thalamocortical Network Atrophy in Sporadic Behavioural Variant Frontotemporal Dementia

**DOI:** 10.1101/2023.03.31.23287956

**Authors:** David Jakabek, Brian D. Power, Nicola Spotorno, Matthew D. Macfarlane, Mark Walterfang, Dennis Velakoulis, Christer Nilsson, Maria Landqvist Waldö, Jimmy Lätt, Markus Nilsson, Danielle van Westen, Olof Lindberg, Jeffrey C. L. Looi, Alexander F. Santillo

## Abstract

Using multi-block methods we combined multimodal neuroimaging metrics of thalamic morphology, thalamic white matter tract diffusion metrics, and cortical thickness to examine changes in behavioural variant frontotemporal dementia. (bvFTD). Twenty-three patients with sporadic bvFTD and 24 healthy controls underwent structural and diffusion MRI scans. Clinical severity was assessed using the Clinical Dementia Rating scale and behavioural severity using the Frontal Behaviour Inventory by patient caregivers. Thalamic volumes were manually segmented. Anterior and posterior thalamic radiation fractional anisotropy and mean diffusivity were extracted using Tract-Based Spatial Statistics. Finally, cortical thickness was assessed using Freesurfer. We used shape analyses, diffusion measures, and cortical thickness as features in sparse multi-block partial least squares (PLS) discriminatory analyses to classify participants within bvFTD or healthy control groups. Sparsity was tuned with five-fold cross-validation repeated 10 times. Final model fit was assessed using permutation testing. Additionally, sparse multi-block PLS was used to examine associations between imaging features and measures of dementia severity. The main features predicting bvFTD group membership were bilateral anterior-dorsal thalamic atrophy, increase in mean diffusivity of thalamic projections, and frontotemporal cortical thinning. The model had a sensitivity of 96%, specificity of 68%, and was statistically significant using permutation testing (p = 0.012). For measures of dementia severity, we found similar involvement of regional thalamic and cortical areas as in discrimination analyses, although more extensive thalamo-cortical white matter metric changes. Using multimodal neuroimaging, we demonstrate combined structural network dysfunction of anterior cortical regions, cortical-thalamic projections, and anterior thalamic regions in sporadic bvFTD.

## 1. Introduction

Behavioural variant frontotemporal dementia (bvFTD) is a common cause of younger onset dementia and is characterised by alterations to personality, cognition, and behaviour. Most cases are sporadic, although genetic mutations (e.g., microtubule-associated protein tau, *MAPT*; progranulin, *GRN*; and chromosome 9 open reading frame 72, *C9ORF72*) have been identified in approximately 20% of cases (Blauwendraat et al., 2018; Wagner et al., 2021). While bvFTD is characterised clinically by frontal and temporal lobe cortical atrophy (Rascovsky et al., 2011), there is also degeneration of subcortical brain structures (Brettschneider et al., 2014; Hornberger et al., 2012). The relative contributions of subcortical and cortical atrophy to the clinical presentation remain unclear. Our interest is in bvFTD as a failure of subcortical-cortical networks (Looi et al., 2014), possibly through a prion-like spread of pathology (Hock & Polymenidou, 2016). From a network perspective, the thalamus has known anatomical projections that modulate and integrate signals between diverse cortical and subcortical areas (Hwang et al., 2017; Sherman, 2007). Thus, the thalamus is of strategic interest as a key hub in a potential network neuropathological model of bvFTD.

In bvFTD, thalamic structural atrophy, reduced white matter connectivity, and abnormal functional connectivity has been demonstrated in neuroimaging and neuropathological studies in both sporadic and genetic bvFTD cases. Patients with *C9ORF72* genotypes had more histological thalamic degeneration compared to sporadic bvFTD (Yang et al., 2017), and FDG-PET studies demonstrated significant thalamic hypometabolism in *C9ORF72* bvFTD compared to sporadic bvFTD (Diehl-Schmid et al., 2019). Other research using MRI segmentation has demonstrated that while thalamic atrophy is maximal in *C9ORF72* genotype (Cash et al., 2018), thalamic atrophy is present in all bvFTD genetic groups (Bocchetta et al., 2018). In diffusion imaging studies, the anterior thalamic radiations have repeatedly demonstrated decreased fractional anisotropy (FA) and increased mean diffusivity (MD) in bvFTD (Daianu et al., 2016; Mahoney et al., 2014; Möller et al., 2015; Zhang et al., 2009), and these diffusion measures are associated with the degree of executive impairment (Tartaglia et al., 2012). A functional network study of bvFTD found reduced connectivity within the salience network, localised to the medial pulvinar of the thalamus (Lee et al., 2014). Thus, atrophy of the thalamus, reduced thalamic structural connectivity, and reduced functional corticothalamic connectivity are observed in bvFTD.

Although these studies mostly suggest global thalamic atrophy (having examined the thalamus as a homogeneous neural structure), the thalamus, being composed of multiple nuclei each associated a diverse range of neural functions, should be rather be considered and explored as a heterogeneous structure, particularly given the varying phenotype in bvFTD (Power & Looi, 2015). Changes in regional thalamic morphology may be observed as a marker of neurodegenerative changes in specific thalamic nuclei. Analogously, our understanding of cortical function would be unduly constrained if we were limited to study whole cortical volume as our only outcome variable. Studies addressing regional thalamic atrophy in sporadic bvFTD are limited. A structural study in a cohort of heterogeneous genetic carriers and sporadic cases found atrophy in anterior thalamic regions which preceded symptomatic bvFTD (Cury et al., 2019). This is in keeping with the known connectivity of the anterior parts of the thalamus to frontal lobes (Behrens et al., 2003). However, this contrasts with functional network research which noted medial pulvinar hypometabolism in *C9ORF72* carriers (Lee et al., 2014). Our previous study, using probabilistic diffusion segmentation of the thalamus, demonstrated atrophy of regions connected to the dorsolateral prefrontal cortex, contrasting with hypertrophy of thalamic regions connected to the medial frontal cortex (Jakabek et al., 2018).

Regional thalamic atrophy can be assessed using statistical shape analyses. However, shape analyses share many of the broader statistical challenges of neuroimaging data, including relatively small sample sizes for the thousands of imaging elements which form dependent variables. Mass univariate testing with correction for multiple comparisons (such as false-discovery rate (FDR) correction) is commonly employed in shape analyses (e.g., spherical harmonic point correspondence framework, SPHARM-PDM (Styner et al., 2006). Alternatively, dimension reduction techniques such as partial least squares (PLS) can be combined with sparse feature selection, discrimination analysis (PLS-DA), and multimodal analyses for integrative analyses across feature sets (Chen et al., 2009). Importantly, the latter multi-block methods allow integration of multimodal datasets (e.g., shape, structural, and diffusion data) to be associated with an outcome variable (e.g., disease status) to better describe neuroimaging phenotypes of diseases (e.g., Avants et al., 2014, 2021; Hoagey et al., 2019; McIntosh & Lobaugh, 2004).

From a methodological perspective we aim to apply shape analysis within a multi-block sparse partial least squares analysis strategy to allow us to simultaneously approach three facets of thalamic degeneration: thalamic atrophy via shape change, frontal-thalamic degeneration via altered diffuse tensor imaging metrics, and cortical atrophy. To our knowledge, this approach, which allows more detailed mapping of the structural integrity of corticothalamic circuitry, has not been applied previously. Secondarily, from a clinical perspective, we aim to describe the involvement of thalamic subregions in sporadic bvFTD and the relationship of subregional change to cortical, tractographic and clinical features. We hypothesise that:

1. Patients with sporadic bvFTD, compared to healthy controls, will have thalamic volumetric atrophy which is predominantly driven by anterior and pulvinar regions in shape analyses; and thalamic-cortical tracts will show reduced fractional anisotropy (FA) and increased mean diffusivity (MD), and;
2. Cortical, shape, and white matter changes will be correlated across known corresponding cortical-subcortical regions and pathways, and;
3. The extent of volume, shape, and diffusion measure changes in sporadic bvFTD will relate to the severity of clinical measures.

## 2. Materials and methods

### 2.1 Participants

Participants in this study were enrolled in the Lund Prospective Frontotemporal Dementia Study (LUPROFS) (see Santillo et al., 2013). The diagnosis of bvFTD was made according to International bvFTD Consortium Criteria (Rascovsky et al., 2011). Assessments included clinical interview, informant history with ratings of behavioural disturbances and disease severity, clinical examination, neuropsychological assessment (outlined below for bvFTD participants), brain MRI (structural and diffusion imaging), genetic testing, and cerebrospinal fluid analysis of amyloid-beta-42, total tau, and phosphorylated tau (indicators of Alzheimer’s Disease). Healthy controls underwent a similar assessment but did not undergo genetic testing. Genetic screening for expansions/mutations in the genes of *C9ORF72*, *MAPT*, and *GRN* was performed in all patients. We excluded three patients with genetic variant bvFTD to focus on sporadic bvFTD. Post-mortem neuropathological examination was performed for a limited number of patients. Two patients and two control participants did not undergo diffusion imaging and so were excluded. Ethical approval was obtained from the Regional Ethical Review Board, Lund, Sweden, and the Australian National University. Issues around obtaining informed consent in the LUPROFS cohort have been outlined previously (see Macfarlane et al., 2015).

### 2.2 Clinical severity and behavioural ratings

For disease severity the LUPROFS study utilised the Frontotemporal Lobar modified Clinical Dementia Rating (FLTD-CDR) score (Knopman et al., 2008). We utilised the “sum of boxes” aggregate score (FTLD-CDR-SB) for subsequent analysis as it is more normally distributed. For quantification of behavioural disturbances the Frontal Behavioural Inventory (FBI) was utilised (Kertesz et al., 1997). The FBI rates 24 behaviours on a 0-3 scale (with higher scores reflecting more severe disturbance); FBI items 1-10 represent negative symptoms (e.g., apathy and neglect), and FBI items 12-22 represent positive symptoms (e.g., impulsivity and hyperorality), and reporting these composites FBI^1-10^, FBI^12-22^) is common in bvFTD studies (Kertesz et al., 1997; Macfarlane et al., 2015; Zamboni et al., 2008). Participants with bvFTD underwent both bedside cognitive and full neuropsychological testing. However, due to the small number of participants who completed all subscales without floor effects, the neuropsychological tests were not analysed further.

### 2.3 Magnetic Resonance Imaging

MRI was performed using a 3.0T Philips MR scanner, with an eight-channel head coil (Philips Achieva®, Philips Medical Systems, Best, The Netherlands). High resolution anatomical images were acquired using a T1-weighted turbo field echo pulse sequence (TR 8ms; TE 4ms; T1 650 ms; FA 10°; NEX2; SENSE-factor 2.4; matrix 240×240; FOV 240×240 mm^2^; resulting voxel size 1×1×1 mm^3^); 175 contiguous coronal slices were obtained. Diffusion weighted imaging (DWI) was performed with an echo-planar single-shot spin echo sequence. Diffusion encoding was performed in 48 directions at b-value = 800 s/mm^2^, one b-value=0 volume, isotropic voxel size 2×2×2 mm^3^, TR 7881 ms, and TE 90 ms.

### 2.4 Thalamic segmentation

Boundary tracing was performed by an experienced investigator (BDP) who had previously developed a protocol for manual segmentation of the dorsal thalamus on T1 MRI scans (Power et al., 2015) using ANALYZE 11.0 software (Mayo Biomedical Imaging Resource, Rochester, Minnesota, USA). The investigator was blind to the clinical status of participants while conducting manual segmentation analysis. Reliability of image analysis was performed by measuring intra-class correlations (type A intra-class correlation coefficients using an absolute agreement definition; scores 0.95 and 0.98) between initial segmentation and random repeated segmentation of both left and right thalami of 5 subjects (a total of 10 measurements) (Power et al., 2015), calculated in SPSS 17.0 (IBM Corporation, Somers, New York, USA).

### 2.5 Shape analysis

Manual thalamic segmentations were pre-processed by iterative rigid alignment to a study-specific thalamic template using the advanced normalisation tools package (Avants et al., 2008) and subsequently, 3D models were constructed of the aligned segmentations using the marching cubes algorithm. Shape processing of the thalamus was conducted using the Deformetrica 4.0.3 software which utilises large deformation diffeomorphic metric mapping (Bône et al., 2018; Durrleman et al., 2014). Briefly, the deformation from the template model to each participant’s model was described using deformation vectors (“momenta”) in 3D space (object kernel width = 3, noise = 3, and deformation kernel = 5) yielding 450 momenta for each shape. These momenta were utilised in the subsequent PLS family of analyses. Momenta which were selected following sparsity analyses (see below) were then projected from the template shape. For visualisation purposes, the template shape was deformed according to PLS loadings, and the displacement along the vertex normals between the deformed shape and the template shape are displayed. Lastly, we compared this approach against the SPHARM-PDM shape analysis methodology as detailed in the supplemental information.

### 2.6 Cortical analyses

T1 scans were processed using Freesurfer 6.0 software (http://surfer. nmr.mgh.harvard.edu). Briefly, grey, white, and cerebrospinal fluid boundaries are automatically determined, and parcellation was performed using the Desikan–Killiany atlas (Desikan et al., 2006). Cortical thickness was extracted, calculated as the closest distance from the gray/white boundary to the gray/CSF boundary at each vertex on the surface (Fischl & Dale, 2000).

### 2.7 DTI analyses

The DWI data were corrected for motion and eddy current induced artifacts using an extrapolation-based registration approach (Nilsson et al., 2015). The diffusion tensor model was subsequently fitted using FMRIB Software Library to compute FA and MD. Tract-based spatial statistics pipeline (Smith et al., 2006) was then applied. In brief, the FA maps were warped to the FMRIB58_FA standard template using FLS’s non-linear registration tool. All the warped FA maps were subsequently averaged to create a mean FA template, from which the FA skeleton was computed. Bothe the FA and the MD maps of each participant were then projected onto the skeleton. The anterior and posterior thalamic radiations were defined using the Johns Hopkins University white matter atlas (JHU, available in FSL (Hua et al., 2008). The mask of the tract of interest were intercepted with study-wise FA skeleton and the median values of FA, MD were extracted from each tract for each participant.

### 2.8 Statistical analyses

All analyses were conducted in R version 3.6 and for the PLS family of analyses we used the mixOmics package (Rohart et al., 2017; Singh et al., 2019). Comparisons of demographic data, cortical thickness, and DTI data were conducted using linear models, and for gender, using Pearson Chi-square. For shape analyses data were controlled for intracranial volume and age, whilst analyses of DTI and and cortical thickness measures were controlled for age only. To control for covariates in PLS analyses, covariates were regressed out from relevant measures and the regression residuals used for subsequent analyses. We utilised Bonferroni-like corrections for analyses of DTI data (significant p < 0.012, 0.05 corrected for 4 tracts) and cortical regions (significant p < 0.001, 0.05 corrected for 34 parcellations and expecting non-independence between hemispheres).

Our primary analysis involved sparse multi-block partial least squares analysis (multi-block sPLS). For all analyses we combined left and right hemisphere data in each block. We used discrimination analysis for the categorial outcome of healthy control vs sporadic bvFTD classification, and regression models for the behavioural measures of bvFTD (i.e., FTLD-CDR-SB, FBI scores, and FBI subscales). Given the modest sample size we used nested-cross validation for sparsity tuning to reduce over-fitting and improve generalisation (Cawley & Talbot, 2010). We used 5-fold cross validation in the outer loop over 10 repeats, and 4-fold cross validation repeated 20 times for the inner loop, across all permutations of shape, cortical, and diffusion measures. We selected the sparsity parameters with the lowest balanced error rate of the centroids distance (for the categorial measure) or mean square error (for continuous measures). One principal component was specified in all analyses. Final model significance was determined by permutation of either group labels or behavioural scores using 1000 repeats. Significance for PLS analyses was p < 0.05 and indicates that the final model predictive ability of group membership or behavioural scores is less likely to occur by chance alone. Our R code for all shape and PLS analyses are publicly available at: https://github.com/djakabek/multimodal

Additionally, validation of this multi-block sPLS approach was conducted using standard SPHARM analysis pipelines and non-sparse partial least squares analysis. These validation analyses are detailed in the supplemental material.

## 3. Results

### 3.1 Sample demographics

Demographic, clinical, and thalamus structural data of patients included in the study are presented in Table 1. Of the 23 eligible participants with bvFTD, 17 had probable FTD, and 6 had definite bvFTD according to consortium criteria. Participants with bvFTD did not statistically differ from healthy controls on age or gender (p > 0.3), although did have fewer years of education (p = 0.012).

**Table 1.** Participant characteristics and structural data

| Demographics | Control |  | bvFTD |  |
| --- | --- | --- | --- | --- |
| n | 24 |  | 23 |  |
| Male | 11 |  | 15 |  |
|  | M | SD | M | SD |
| Age | 66 | 11 | 69 | 8 |
| Education (years) | 12 | 3 | 10 | 3 |
| MMSE | 29* | 1 | 22 | 5 |
| FTLD-CDR-SB |  |  | 8.7 | 3.8 |
| FBI Total score |  |  | 26.2 | 8.2 |
| FBI 1-10 score |  |  | 16.0 | 5.5 |
| FBI 12-22 score |  |  | 9.2 | 4.7 |
| Left thalamus (mm<sup>3</sup>) | 4714 | 644 | 4083 | 724 |
| Right thalamus (mm<sup>3</sup>) | 4681 | 644 | 4224 | 737 |

Abbreviations: bvFTD = Behavioural Variant Frontotemporal Dementia; FTLD- CDR-SB = Frontotemporal lobar degeneration clinical dementia rating tool sum of boxes score; FBI = Frontal Behavioural Inventory; MMSE = Mini Mental state examination. \* MMSE available for 13 participants.

### 3.2 Group comparisons

The total left thalamus was significantly smaller in volume in patients with bvFTD compared to controls (Beta = −757, 95% CI −757 to −89, p = 0.014). The right thalamus was also smaller in patients with bvFTD compared to controls however this was not statistically significant (Beta = −247, 95% CI −585 to 91, p = 0.147). Diffusion measures between groups are shown in Table 2 and demonstrate that in bvFTD there is significantly lower FA and higher MD for both anterior and posterior thalamic radiations. Lastly, significant cortical thinning was observed in bilateral frontal regions (coefficients provided in supplemental table 1).

**Table 2.** Tract-based spatial statistics results

| Tract | Control |  | bvFTD |  | Beta | SE | p-value |
| --- | --- | --- | --- | --- | --- | --- | --- |
|  | M | SE | M | SE |  |  |  |
| FA |  |  |  |  |  |  |  |
| Left ATR | 0.497 | 0.012 | 0.440 | 0.012 | -0.057 | 0.017 | 0.002 |
| Right ATR | 0.491 | 0.012 | 0.426 | 0.013 | -0.065 | 0.018 | 0.001 |
| Left PTR | 0.453 | 0.009 | 0.429 | 0.010 | -0.024 | 0.014 | 0.082 |
| Right PTR | 0.453 | 0.009 | 0.429 | 0.010 | -0.024 | 0.014 | 0.082 |
| MD ($\times 10^{-4}$) | | | | | | | |
| Left ATR | 0.761 | 0.020 | 0.889 | 0.020 | 0.128 | 0.028 | <0.001 |
| Right ATR | 0.743 | 0.019 | 0.849 | 0.020 | 0.106 | 0.028 | <0.001 |
| Left PTR | 0.835 | 0.017 | 0.896 | 0.018 | 0.061 | 0.025 | 0.018 |
| Right PTR | 0.841 | 0.018 | 0.931 | 0.019 | 0.090 | 0.026 | 0.001 |

4 Estimated marginal means shown adjusted for age. M= mean, SE= Standard Error of 5 the Mean. FA = fractional anisotropy, MD = mean diffusivity, ATR = anterior 6 thalamic radiation, PTR = posterior thalamic radiation.

Group differences using multi-block sPLS-DA are displayed in Figure 1. Sparsity tuning selected 480 thalamic deformation momenta, 56 cortical regions, and 4 DTI measures across both hemispheres. In the bvFTD patient group there was reduced bilateral cortical thickness in anterior regions (orbitofrontal, dorsolateral, and medial prefrontal) and the left insula. There was anterior-dorsal deflation in bilateral thalami, and deflation in the left mediodorsal region. An increase in MD for the bilateral anterior thalamic radiations had the highest weight in discriminating between healthy controls from patients with bvFTD, and reduced FA of the right anterior thalamic radiation and increased MD of the right posterior thalamic radiation were also found to discriminate between groups. This model misclassified one healthy control and seven bvFTD patients (sensitivity = 96%, specificity = 68%) and was statistically significant using permutation testing (p = 0.019).

**Figure 1.**
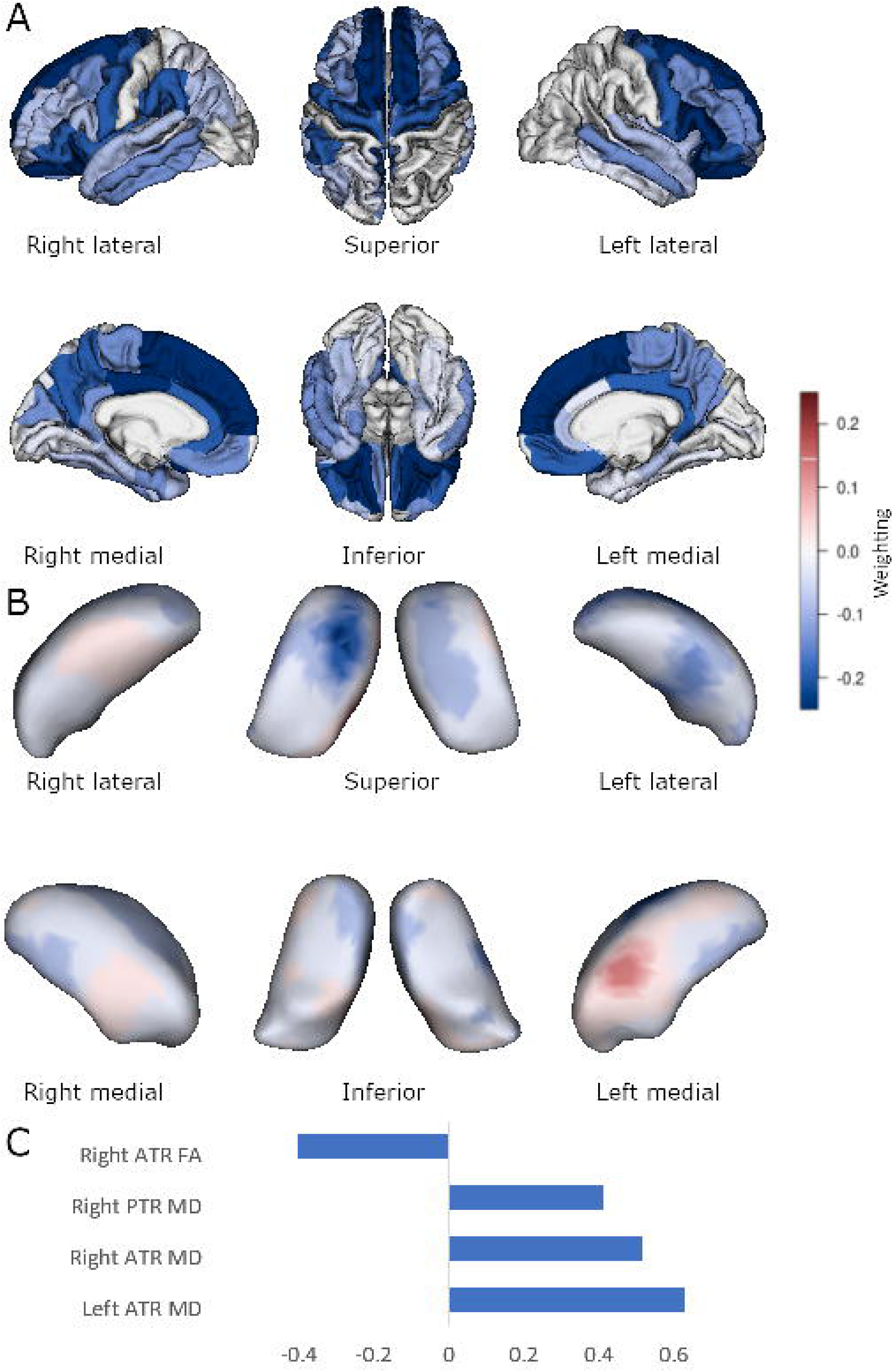
Sparse PLS-DA group comparison. Multiblock group comparison. Panel A shows cortical loading values for the sparse model. Panel B shows displacement from an average thalamic shape after deformation by sparsely selected momenta. Panel C shows sparsely selected DTI tracts. ATR, anterior thalamic radiation; PTR, posterior thalamic radiation; FA, fractional anisotropy; MD, mean diffusivity. Scales show relative weighting in the discrimination selection model and are comparable in colour between Panels A and B.

### 3.3 Clinical severity and behavioural measures

Using volumetric measures and linear regression, there was significant reductions in total thalamic volumes associated with FTLD-CDR-SB scores (left Beta = −105, SE = 30, p=.002; right Beta = −78, SE = 34, p = 0.031), the FBI total score (left Beta = −55, SE = 13, p = 0.001; right Beta = −35, SE = 15, p = 0.037), and the FBI 1-10 score (left Beta = −88, SE = 14, p < 0.001, right Beta = −52, SE = 21, p = 0.024). The FBI 12-20 score was not significant associated with either thalamus volume (p > 0.255). No significant associations were observed for regional cortical or DTI with clinical severity or behavioural measures using a corrected p-value threshold (coefficients provided in supplemental information).

By contrast, significant cortical, shape, and diffusion associations were observed using multi-block sPLS methodology. For the FTLD-CDR-SB score model (shown in Figure 2) there were 56 cortical regions, 510 momenta directions, and all DTI tracts selected, r = 0.74, p = 0.01. Similar significant findings were for the FBI total score (Figure 3, 46 cortical regions, 190 thalamic momenta, all DTI tracts, r = 0.69, p = 0.02) and FBI 1-10 score (Figure 4, 26 cortical regions, 120 thalamic momenta, all DTI tracts, r = 0.69, p = 0.04). The FBI 12-22 subscale model was sparsely reduced to 32 cortical regions, all DTI tracts, and 50 thalamic momenta, but this model was not statistically significant (r = 0.60, p = 0.13).

**Figure 2.**
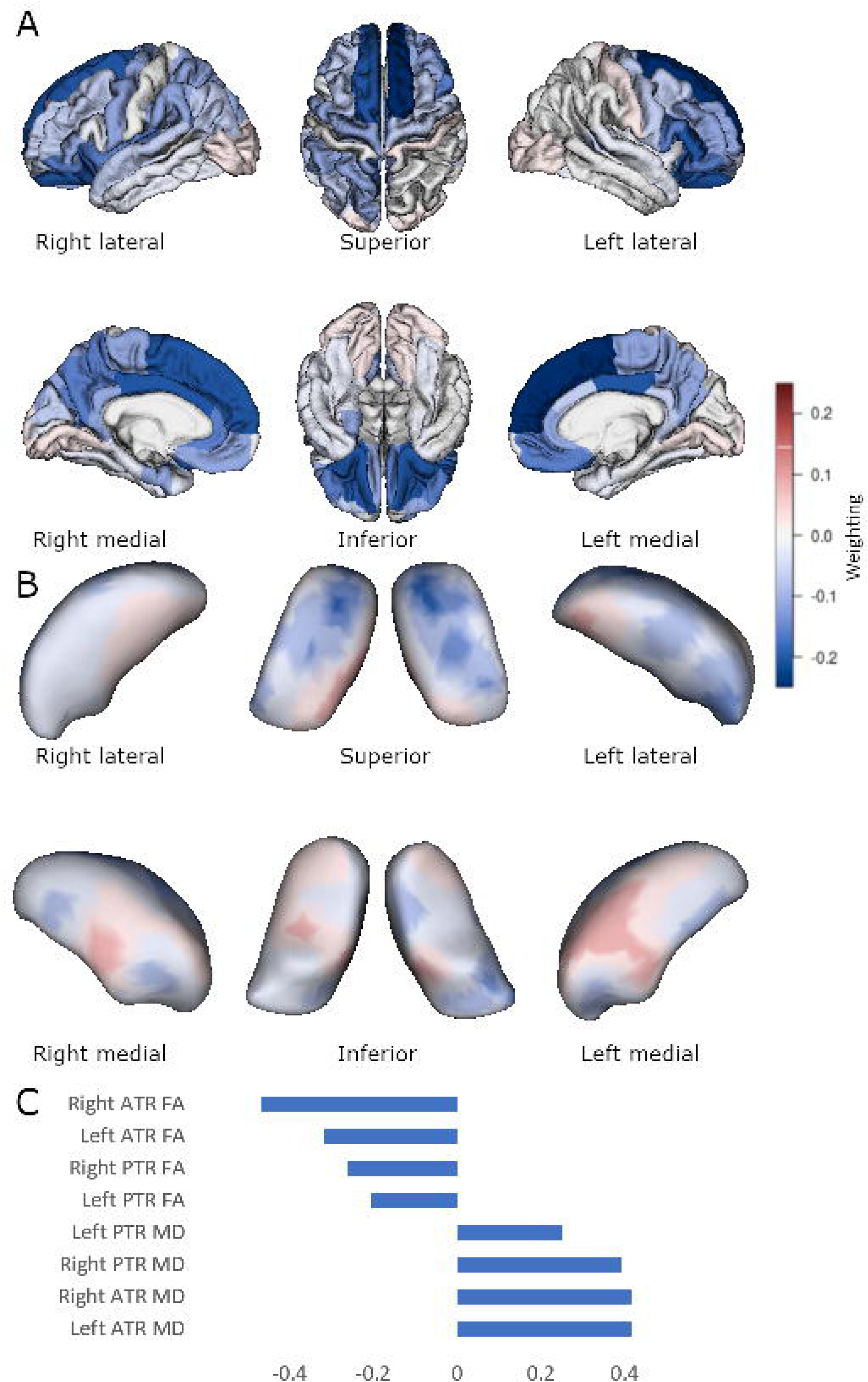
Sparse PLS association with FTLD-CDR-SB. Multiblock group comparison. Panel A shows cortical loading values for the sparse model. Panel B shows displacement from an average thalamic shape after deformation by sparsely selected momenta. Panel C shows sparsely selected DTI tracts. ATR, anterior thalamic radiation; PTR, posterior thalamic radiation; FA, fractional anisotropy; MD, mean diffusivity. Scales show weighting in the discrimination selection model and are comparable in colour between Panels A and B.

**Figure 3.**
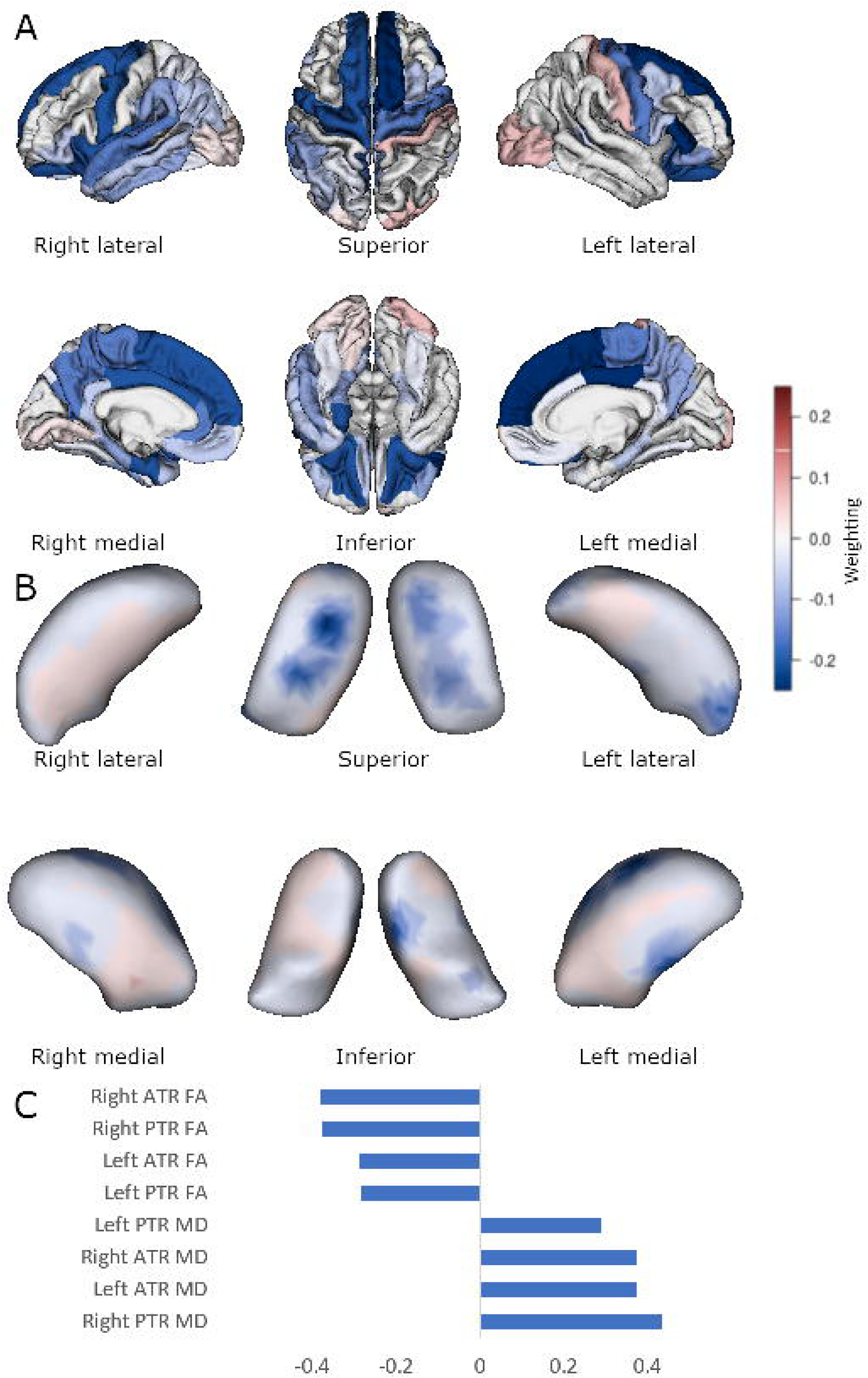
Sparse PLS association with FBI total score. Multiblock group comparison. Panel A shows cortical loading values for the sparse model. Panel B shows displacement from an average thalamic shape after deformation by sparsely selected momenta. Panel C shows sparsely selected DTI tracts. ATR, anterior thalamic radiation; PTR, posterior thalamic radiation; FA, fractional anisotropy; MD, mean diffusivity. Scales show weighting in the discrimination selection model and are comparable in colour between Panels A and B.

**Figure 4.**
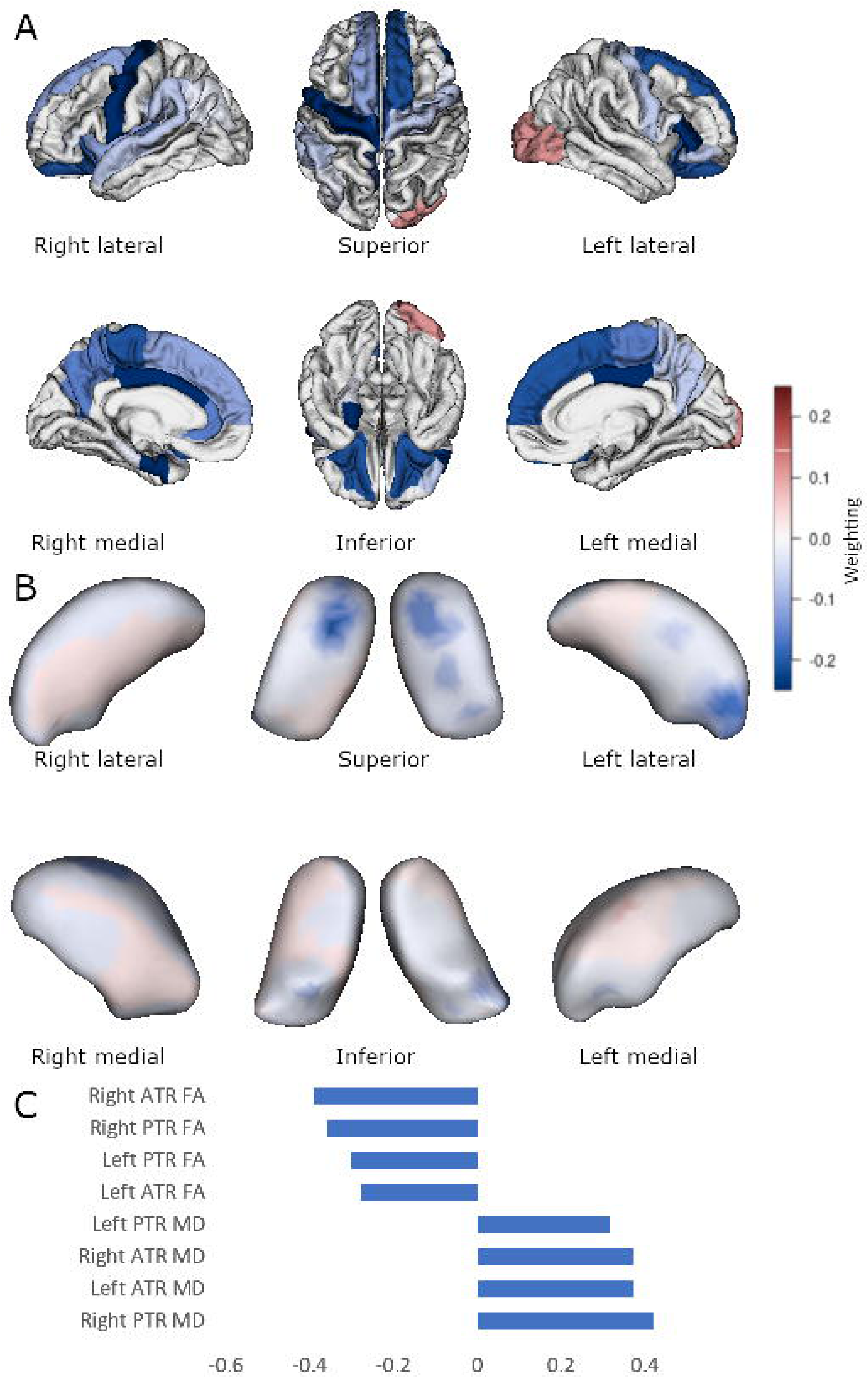
Sparse PLS association with FBI score on items 1 to 10. Multiblock group comparison. Panel A shows cortical loading values for the sparse model. Panel B shows displacement from an average thalamic shape after deformation by sparsely selected momenta. Panel C shows sparsely selected DTI tracts. ATR, anterior thalamic radiation; PTR, posterior thalamic radiation; FA, fractional anisotropy; MD, mean diffusivity. Scales show weighting in the discrimination selection model and are comparable in colour between Panels A and B.

## 4. Discussion

Through innovative use of multi-block sparse partial least squares analyses we demonstrated concurrent changes to thalamic morphometry, diffusion measurements, and cortical thickness in sporadic bvFTD. Sporadic bvFTD group membership was associated with a reduction in bilateral anterior cortical thickness, increased mean diffusivity in anterior thalamic radiations, and deflation of the anterior-dorsal aspect of the thalamus, compared to healthy controls. Moreover, similar regions were associated with worsening bvFTD disease severity quantified via clinical dementia rating and behavioural measures.

Methodologically, using multi-block methods, we demonstrate that cortical, diffusion, and shape features can be combined to predict diagnostic group and disease severity. The association of the multimodality structural neuroimaging features with diagnosis and disease severity establishes these features as a potential neuroanatomical basis of a sporadic bvFTD clinical phenotype.

Cortical thinning in anterior and temporal regions in our sample of sporadic bvFTD is expected, given that the diagnoses of bvFTD was made using International bvFTD Consortium Criteria (Rascovsky et al., 2011) which includes frontal and/or temporal cortical volume loss as a criterion.

Diffusion analyses found predominantly increased MD in the anterior thalamic radiations bilaterally for bvFTD group classification, and on the left anterior thalamic radiation for disease severity. This is consistent with the extant literature on group differences and disease severity in bvFTD, where raised MD is commonly observed (Daianu et al., 2016; Mahoney et al., 2014; Möller et al., 2015; Tartaglia et al., 2012; Zhang et al., 2009). Altered DTI metrics in the anterior thalamic radiations are concordant with our shape findings since afferents and efferents from these nuclei traverse these pathways to the prefrontal cortex. Previously, Schönecker et al. (2018), using volumetric divisions of the thalamus, demonstrated associations between anterior thalamic atrophy and reduced frontal-thalamic DTI metrics which were more pronounced in those with C9ORF72 mutations than healthy controls. This prior study included a mixed sample of frontotemporal dementia and amyotrophic lateral sclerosis patients. Here, we demonstrate that the relationship between thalamic atrophy and frontal-thalamic DTI metric alterations occur in a sporadic bvFTD cohort as well. Moreover, lower FA and higher MD in all tracts were associated with behavioural measures, whereas only higher MD in the anterior tracts was predicted by bvFTD status. This suggests that clinical manifestations of bvFTD may result from disconnection between the cortex and thalamus.

Although the DTI changes are consistent with previous research, the explanatory pathophysiological mechanisms warrant consideration, particularly given the bidirectional nature of frontal-thalamic connections. Early in bvFTD, cortical supragranular layer inclusions and neuronal loss are most characteristically observed, although inclusions are also present in projections neurons of layer V and VI (Brettschneider et al., 2014), which contain corticothalamic drivers and modulators (Sherman, 2007). Thus, both thalamocortical and corticothalamic networks are affected at the earliest stage of bvFTD. Accordingly, the thalamus appears to be affected early in FTD and cannot be dismissed as a “downstream” phenomenon to cortical involvement. Concordantly, studies examining the Papez circuit in bvFTD have shown that involvement of the thalamic anterior nuclear group is of the same magnitude as that of the anterior cingulate cortex (Hornberger et al., 2012). Despite likely early involvement of the thalamus in the bvFTD disease process, the propagation of neuropathology between the cortex and thalamus remains unknown. Possible mechanisms may include direct white matter pathology, network dysfunction (Palop & Mucke, 2010), or direct or indirect disconnection from either frontal or thalamic regions causing trans-synaptic degeneration (Buren, 1963; Looi & Walterfang, 2013).

There is support for our hypothesis of significant involutional shape change in the thalamus. Volumetrically, we found smaller thalamic volumes bilaterally, although statistically significant differences only on the left. This is broadly consistent with previous volumetric studies (Bocchetta et al., 2018; Cardenas et al., 2007; Garibotto et al., 2011). However, not all studies of sporadic bvFTD have found volumetric thalamic atrophy (Schönecker et al., 2018). Regionally, deflation was observed in the anterior nuclear group (mainly constituted by the anteroventral nucleus), ventral anterior, and the dorsal nuclei (Morel et al., 1997). Previous regional volumetric studies have shown conflicting results in sporadic bvFTD; anterior thalamic volumes have been observed to be reduced (Chow et al., 2008) and also, increased (Bede et al., 2018). Our previous research demonstrated volumetric reduction in dorsolateral-connected thalamic regions but increases in orbitofrontal-connected regions (Jakabek et al., 2018). Prior thalamic shape analyses had examined only genetic bvFTD and found pre-symptomatic (over five years before clinical symptoms) anterior atrophy (Cury et al., 2019). In a comprehensive genetic bvFTD study, automated thalamus parcellation found that the mediodorsal nucleus was the most characteristically affected sub nucleus (Bocchetta et al., 2018), in contrast to our results. Using shape analyses rather than predetermined thalamic sub-nuclear volumetric divisions we demonstrate marked anterior-dorsal atrophy in sporadic bvFTD.

From a methodological perspective, our analysis of thalamic shape involves no *a priori* assumptions on the regions of deformation, and thus may have theoretical advantages over parcellation methods employed in other studies (e.g., Bede et al., 2018; Bocchetta et al., 2018). Moreover, by utilising the multi-block sparse PLS framework, we can combine different imaging modalities across subjects to derive parsimonious correlated predictors of disease or disease severity.

Notably, there was minimal involvement of thalamic sensory or motor nuclei. This suggests that the aberrant processing of tactile sensory information in bvFTD is not attributable to disturbance in this part of the system. Additionally, the pulvinar was not affected in our cohort of sporadic bvFTD. This suggests that pulvinar involvement may be a specific feature of *C9ORF72* bvFTD, in line with other studies (Bocchetta et al., 2018). Furthermore, there were no significant correlations with the FBI 12-22 subscales. This may be attributed to specific dysexecutive behaviour being correlated with atrophy of the striatum (e.g., caudate; Macfarlane et al., 2015), whereas thalamic atrophy may be associated with general cognitive, behavioural and functional decline. Alternatively, our modest sample size may not have had sufficient power to detect such an association. Additionally, given the modest sample size, there is the risk of overfitting using the PLS methodology which we have minimised with the use of nested cross-validation.

In conclusion, we have demonstrated that there is neural network-based thalamic atrophy in sporadic bvFTD, which, in turn, is correlated with tractography and cortical changes. Furthermore, these changes are associated with measures of clinical severity and behavioural disturbance. The characterisation of combination of neuroanatomical and clinical features may form the basis of a more definitive clinical phenotype for sporadic bvFTD, while acknowledging the variable genetic basis of sporadic bvFTD. Anterior and dorsal nuclei of the thalamus seem to be most affected in sporadic bvFTD, whilst ventral and posterior nuclei are affected to a lesser extent. Considering that thalamic function modulates cortico-cortical communication, this work provides a new impetus to further examine thalamic imaging and histopathological changes as potential hallmarks in the spatiotemporal development and progression of bvFTD.

## Data Availability

Anonymized data will be shared by request from a qualified academic investigator for the sole purpose of replicating procedures and results presented in the article if data transfer is in agreement with EU legislation on the general data protection regulation and decisions by the Ethical Review Board of Sweden and Region Skane, which should be regulated in a material transfer agreement. Code is available at https://github.com/djakabek/multimodal

## Abbreviations

bvFTD: behavioural variant frontotemporal dementia
FA: fractional anisotropy
MD: mean diffusivity
FDR: false discovery rate
SPHARM-PDM: spherical harmonic point distribution model
PLS: partial least squares
PLS-DA: partial least squares - discriminant analysis.

## References

Avants, B. B., Epstein, C. L., Grossman, M., & Gee, J. C. (2008). Symmetric diffeomorphic image registration with cross-correlation: Evaluating automated labeling of elderly and neurodegenerative brain. Medical Image Analysis, 12(1), 26–41. https://doi.org/10.1016/j.media.2007.06.004

Avants, B. B., Libon, D. J., Rascovsky, K., Boller, A., McMillan, C. T., Massimo, L., Coslett, H. B., Chatterjee, A., Gross, R. G., & Grossman, M. (2014). Sparse canonical correlation analysis relates network-level atrophy to multivariate cognitive measures in a neurodegenerative population. NeuroImage, 84, 698– 711. https://doi.org/10.1016/j.neuroimage.2013.09.048

Avants, B. B., Tustison, N. J., & Stone, J. R. (2021). Similarity-driven multi-view embeddings from high-dimensional biomedical data. Nature Computational Science, 1(2), 143–152. https://doi.org/10.1038/s43588-021-00029-8

Bede, P., Omer, T., Finegan, E., Chipika, R. H., Iyer, P. M., Doherty, M. A., Vajda, A., Pender, N., McLaughlin, R. L., Hutchinson, S., & Hardiman, O. (2018). Connectivity-based characterisation of subcortical grey matter pathology in frontotemporal dementia and ALS: A multimodal neuroimaging study. Brain Imaging and Behavior, 12(6), 1696–1707. https://doi.org/10.1007/s11682-018-9837-9

Behrens, T. E. J., Johansen-Berg, H., Woolrich, M. W., Smith, S. M., Wheeler-Kingshott, C. a. M., Boulby, P. A., Barker, G. J., Sillery, E. L., Sheehan, K., Ciccarelli, O., Thompson, A. J., Brady, J. M., & Matthews, P. M. (2003). Non-invasive mapping of connections between human thalamus and cortex using diffusion imaging. Nature Neuroscience, 6(7), 750–757. https://doi.org/10.1038/nn1075

Blauwendraat, C., Wilke, C., Simón-Sánchez, J., Jansen, I. E., Reifschneider, A., Capell, A., Haass, C., Castillo-Lizardo, M., Biskup, S., Maetzler, W., Rizzu, P., Heutink, P., & Synofzik, M. (2018). The wide genetic landscape of clinical frontotemporal dementia: Systematic combined sequencing of 121 consecutive subjects. Genetics in Medicine, 20(2), Article 2. https://doi.org/10.1038/gim.2017.102

Bocchetta, M., Gordon, E., Cardoso, M. J., Modat, M., Ourselin, S., Warren, J. D., & Rohrer, J. D. (2018). Thalamic atrophy in frontotemporal dementia—Not just a C9orf72 problem. NeuroImage: Clinical, 18, 675–681. https://doi.org/10.1016/j.nicl.2018.02.019

Bône, A., Louis, M., Martin, B., & Durrleman, S. (2018). Deformetrica 4: An Open-Source Software for Statistical Shape Analysis. In M. Reuter, C. Wachinger, H. Lombaert, B. Paniagua, M. Lüthi, & B. Egger (Eds.), Shape in Medical Imaging (pp. 3–13). Springer International Publishing. https://doi.org/10.1007/978-3-030-04747-4_1

Brettschneider, J., Del Tredici, K., Irwin, D. J., Grossman, M., Robinson, J. L., Toledo, J. B., Fang, L., Van Deerlin, V. M., Ludolph, A. C., Lee, V. M.-Y., Braak, H., & Trojanowski, J. Q. (2014). Sequential distribution of pTDP-43 pathology in behavioral variant frontotemporal dementia (bvFTD). Acta Neuropathologica, 127(3), 423–439. https://doi.org/10.1007/s00401-013-1238-y

Buren, J. M. V. (1963). Trans-synaptic retrograde degeneration in the visual system of primates. Journal of Neurology, Neurosurgery & Psychiatry, 26(5), 402–409. https://doi.org/10.1136/jnnp.26.5.402

Cardenas, V. A., Boxer, A. L., Chao, L. L., Gorno-Tempini, M. L., Miller, B. L., Weiner, M. W., & Studholme, C. (2007). Deformation Morphometry Reveals Brain Atrophy in Frontotemporal Dementia. Archives of Neurology, 64(6), 873–877. https://doi.org/10.1001/archneur.64.6.873

Cash, D. M., Bocchetta, M., Thomas, D. L., Dick, K. M., van Swieten, J. C., Borroni, B., Galimberti, D., Masellis, M., Tartaglia, M. C., Rowe, J. B., Graff, C., Tagliavini, F., Frisoni, G. B., Laforce, R., Finger, E., de Mendonça, A., Sorbi, S., Rossor, M. N., Ourselin, S., … Warren, J. (2018). Patterns of gray matter atrophy in genetic frontotemporal dementia: Results from the GENFI study. Neurobiology of Aging, 62, 191–196. https://doi.org/10.1016/j.neurobiolaging.2017.10.008

Cawley, G. C., & Talbot, N. L. C. (2010). On Over-fitting in Model Selection and Subsequent Selection Bias in Performance Evaluation. Journal of Machine Learning Research, 11(70), 2079–2107.

Chen, K., Reiman, E. M., Huan, Z., Caselli, R. J., Bandy, D., Ayutyanont, N., & Alexander, G. E. (2009). Linking Functional and Structural Brain Images with Multivariate Network Analyses: A Novel Application of the Partial Least Square Method. NeuroImage, 47(2), 602–610. https://doi.org/10.1016/j.neuroimage.2009.04.053

Chow, T. W., Izenberg, A., Binns, M. A., Freedman, M., Stuss, D. T., Scott, C. J. M., Ramirez, J., & Black, S. E. (2008). Magnetic Resonance Imaging in Frontotemporal Dementia Shows Subcortical Atrophy. Dementia and Geriatric Cognitive Disorders, 26(1), 79–88. https://doi.org/10.1159/000144028

Cury, C., Durrleman, S., Cash, D. M., Lorenzi, M., Nicholas, J. M., Bocchetta, M., van Swieten, J. C., Borroni, B., Galimberti, D., Masellis, M., Tartaglia, M. C., Rowe, J. B., Graff, C., Tagliavini, F., Frisoni, G. B., Laforce, R., Finger, E., de Mendonça, A., Sorbi, S., … Warren, J. (2019). Spatiotemporal analysis for detection of pre-symptomatic shape changes in neurodegenerative diseases: Initial application to the GENFI cohort. NeuroImage, 188, 282–290. https://doi.org/10.1016/j.neuroimage.2018.11.063

Daianu, M., Mendez, M. F., Baboyan, V. G., Jin, Y., Melrose, R. J., Jimenez, E. E., & Thompson, P. M. (2016). An advanced white matter tract analysis in frontotemporal dementia and early-onset Alzheimer’s disease. Brain Imaging and Behavior, 10(4), 1038–1053. https://doi.org/10.1007/s11682-015-9458-5

Diehl-Schmid, J., Licata, A., Goldhardt, O., Förstl, H., Yakushew, I., Otto, M., Anderl-Straub, S., Beer, A., Ludolph, A. C., Landwehrmeyer, G. B., Levin, J., Danek, A., Fliessbach, K., Spottke, A., Fassbender, K., Lyros, E., Prudlo, J., Krause, B. J., Volk, A., … Grimmer, T. (2019). FDG-PET underscores the key role of the thalamus in frontotemporal lobar degeneration caused by C9ORF72 mutations. Translational Psychiatry, 9(1), 1–11. https://doi.org/10.1038/s41398-019-0381-1

Durrleman, S., Prastawa, M., Charon, N., Korenberg, J. R., Joshi, S., Gerig, G., & Trouvé, A. (2014). Morphometry of anatomical shape complexes with dense deformations and sparse parameters. NeuroImage, 101, 35–49. https://doi.org/10.1016/j.neuroimage.2014.06.043

Fischl, B., & Dale, A. M. (2000). Measuring the thickness of the human cerebral cortex from magnetic resonance images. Proceedings of the National Academy of Sciences, 97(20), 11050–11055. https://doi.org/10.1073/pnas.200033797

Garibotto, V., Borroni, B., Agosti, C., Premi, E., Alberici, A., Eickhoff, S. B., Brambati, S. M., Bellelli, G., Gasparotti, R., Perani, D., & Padovani, A. (2011). Subcortical and deep cortical atrophy in Frontotemporal Lobar Degeneration. Neurobiology of Aging, 32(5), 875–884. https://doi.org/10.1016/j.neurobiolaging.2009.05.004

Hoagey, D. A., Rieck, J. R., Rodrigue, K. M., & Kennedy, K. M. (2019). Joint contributions of cortical morphometry and white matter microstructure in healthy brain aging: A partial least squares correlation analysis. Human Brain Mapping, 40(18), 5315–5329. https://doi.org/10.1002/hbm.24774

Hock, E.-M., & Polymenidou, M. (2016). Prion-like propagation as a pathogenic principle in frontotemporal dementia. Journal of Neurochemistry, 138, 163– 183. https://doi.org/10.1111/jnc.13668

Hornberger, M., Wong, S., Tan, R., Irish, M., Piguet, O., Kril, J., Hodges, J. R., & Halliday, G. (2012). In vivo and post-mortem memory circuit integrity in frontotemporal dementia and Alzheimer’s disease. Brain, 135(10), 3015– 3025. https://doi.org/10.1093/brain/aws239

Hua, K., Zhang, J., Wakana, S., Jiang, H., Li, X., Reich, D. S., Calabresi, P. A., Pekar, J. J., van Zijl, P. C. M., & Mori, S. (2008). Tract Probability Maps in Stereotaxic Spaces: Analyses of White Matter Anatomy and Tract-Specific Quantification. NeuroImage, 39(1), 336–347. https://doi.org/10.1016/j.neuroimage.2007.07.053

Hwang, K., Bertolero, M. A., Liu, W. B., & D’Esposito, M. (2017). The Human Thalamus Is an Integrative Hub for Functional Brain Networks. The Journal of Neuroscience, 37(23), 5594–5607. https://doi.org/10.1523/JNEUROSCI.0067-17.2017

Jakabek, D., Power, B. D., Macfarlane, M. D., Walterfang, M., Velakoulis, D., Westen, D. van, Lätt, J., Nilsson, M., Looi, J. C. L., & Santillo, A. F. (2018). Regional structural hypo- and hyperconnectivity of frontal–striatal and frontal–thalamic pathways in behavioral variant frontotemporal dementia. Human Brain Mapping, 39(10), 4083–4093. https://doi.org/10.1002/hbm.24233

Kertesz, A., Davidson, W., & Fox, H. (1997). Frontal behavioral inventory: Diagnostic criteria for frontal lobe dementia. The Canadian Journal of Neurological Sciences. Le Journal Canadien Des Sciences Neurologiques, 24(1), 29–36.

Knopman, D. S., Kramer, J. H., Boeve, B. F., Caselli, R. J., Graff-Radford, N. R., Mendez, M. F., Miller, B. L., & Mercaldo, N. (2008). Development of methodology for conducting clinical trials in frontotemporal lobar degeneration. Brain, 131(11), 2957–2968. https://doi.org/10.1093/brain/awn234

Lee, S. E., Khazenzon, A. M., Trujillo, A. J., Guo, C. C., Yokoyama, J. S., Sha, S. J., Takada, L. T., Karydas, A. M., Block, N. R., Coppola, G., Pribadi, M., Geschwind, D. H., Rademakers, R., Fong, J. C., Weiner, M. W., Boxer, A. L., Kramer, J. H., Rosen, H. J., Miller, B. L., & Seeley, W. W. (2014). Altered network connectivity in frontotemporal dementia with C9orf72 hexanucleotide repeat expansion. Brain, 137(11), 3047–3060. https://doi.org/10.1093/brain/awu248

Looi, J. C. L., & Walterfang, M. (2013). Striatal morphology as a biomarker in neurodegenerative disease. Molecular Psychiatry, 18(4), 417–424. https://doi.org/10.1038/mp.2012.54

Looi, J. C. L., Walterfang, M., Nilsson, C., Power, B. D., van Westen, D., Velakoulis, D., Wahlund, L.-O., & Thompson, P. M. (2014). The subcortical connectome: Hubs, spokes and the space between - a vision for further research in neurodegenerative disease. The Australian and New Zealand Journal of Psychiatry, 48(4), 306–309. https://doi.org/10.1177/0004867413506753

Macfarlane, M. D., Jakabek, D., Walterfang, M., Vestberg, S., Velakoulis, D., Wilkes, F. A., Nilsson, C., van Westen, D., Looi, J. C. L., & Santillo, A. F. (2015). Striatal Atrophy in the Behavioural Variant of Frontotemporal Dementia: Correlation with Diagnosis, Negative Symptoms and Disease Severity. PloS One, 10(6), e0129692. https://doi.org/10.1371/journal.pone.0129692

Mahoney, C. J., Ridgway, G. R., Malone, I. B., Downey, L. E., Beck, J., Kinnunen, K. M., Schmitz, N., Golden, H. L., Rohrer, J. D., Schott, J. M., Rossor, M. N., Ourselin, S., Mead, S., Fox, N. C., & Warren, J. D. (2014). Profiles of white matter tract pathology in frontotemporal dementia. Human Brain Mapping, 35(8), 4163–4179. https://doi.org/10.1002/hbm.22468

McIntosh, A. R., & Lobaugh, N. J. (2004). Partial least squares analysis of neuroimaging data: Applications and advances. NeuroImage, 23, S250–S263. https://doi.org/10.1016/j.neuroimage.2004.07.020

Möller, C., Hafkemeijer, A., Pijnenburg, Y. A. L., Rombouts, S. A. R. B., van der Grond, J., Dopper, E., van Swieten, J., Versteeg, A., Pouwels, P. J. W., Barkhof, F., Scheltens, P., Vrenken, H., & van der Flier, W. M. (2015). Joint assessment of white matter integrity, cortical and subcortical atrophy to distinguish AD from behavioral variant FTD: A two-center study. NeuroImage: Clinical, 9, 418–429. https://doi.org/10.1016/j.nicl.2015.08.022

Morel, A., Magnin, M., & Jeanmonod, D. (1997). Multiarchitectonic and stereotactic atlas of the human thalamus. Journal of Comparative Neurology, 387(4), 588– 630. https://doi.org/10.1002/(SICI)1096-9861(19971103)387:4<588::AID-CNE8>3.0.CO;2-Z

Nilsson, M., Szczepankiewicz, F., Westen, D. van, & Hansson, O. (2015). Extrapolation-Based References Improve Motion and Eddy-Current Correction of High B-Value DWI Data: Application in Parkinson’s Disease Dementia. PLOS ONE, 10(11), e0141825. https://doi.org/10.1371/journal.pone.0141825

Palop, J. J., & Mucke, L. (2010). Synaptic Depression and Aberrant Excitatory Network Activity in Alzheimer’s Disease: Two Faces of the Same Coin? Neuromolecular Medicine, 12(1), 48–55. https://doi.org/10.1007/s12017-009-8097-7

Power, B. D., & Looi, J. C. (2015). The thalamus as a putative biomarker in neurodegenerative disorders. Australian & New Zealand Journal of Psychiatry, 49(6), 502–518. https://doi.org/10.1177/0004867415585857

Power, B. D., Wilkes, F. A., Hunter-Dickson, M., van Westen, D., Santillo, A. F., Walterfang, M., Nilsson, C., Velakoulis, D., & J. C. L. Looi. (2015). Validation of a protocol for manual segmentation of the thalamus on magnetic resonance imaging scans. Psychiatry Research, 232(1), 98–105. https://doi.org/10.1016/j.pscychresns.2015.02.001

Rascovsky, K., Hodges, J. R., Knopman, D., Mendez, M. F., Kramer, J. H., Neuhaus, J., Swieten, J. C. van, Seelaar, H., Dopper, E. G. P., Onyike, C. U., Hillis, A. E., Josephs, K. A., Boeve, B. F., Kertesz, A., Seeley, W. W., Rankin, K. P., Johnson, J. K., Gorno-Tempini, M.-L., Rosen, H., … Miller, B. L. (2011). Sensitivity of revised diagnostic criteria for the behavioural variant of frontotemporal dementia. Brain, 134(9), 2456–2477. https://doi.org/10.1093/brain/awr179

Rohart, F., Gautier, B., Singh, A., & Cao, K.-A. L. (2017). mixOmics: An R package for ‘omics feature selection and multiple data integration. PLOS Computational Biology, 13(11), e1005752. https://doi.org/10.1371/journal.pcbi.1005752

Santillo, A. F., Mårtensson, J., Lindberg, O., Nilsson, M., Manzouri, A., Waldö, M. L., Westen, D. van, Wahlund, L.-O., Lätt, J., & Nilsson, C. (2013). Diffusion Tensor Tractography versus Volumetric Imaging in the Diagnosis of Behavioral Variant Frontotemporal Dementia. PLOS ONE, 8(7), e66932. https://doi.org/10.1371/journal.pone.0066932

Schönecker, S., Neuhofer, C., Otto, M., Ludolph, A., Kassubek, J., Landwehrmeyer, B., Anderl-Straub, S., Semler, E., Diehl-Schmid, J., Prix, C., Vollmar, C., Fortea, J., Huppertz, H.-J., Arzberger, T., Edbauer, D., Feddersen, B., Dieterich, M., Schroeter, M. L., Volk, A. E., … Levin, J. (2018). Atrophy in the Thalamus But Not Cerebellum Is Specific for C9orf72 FTD and ALS Patients – An Atlas-Based Volumetric MRI Study. Frontiers in Aging Neuroscience, 10, 45. https://doi.org/10.3389/fnagi.2018.00045

Sherman, S. M. (2007). The thalamus is more than just a relay. Current Opinion in Neurobiology, 17(4), 417–422. https://doi.org/10.1016/j.conb.2007.07.003

Singh, A., Shannon, C. P., Gautier, B., Rohart, F., Vacher, M., Tebbutt, S. J., & Lê Cao, K.-A. (2019). DIABLO: An integrative approach for identifying key molecular drivers from multi-omics assays. Bioinformatics, 35(17), 3055– 3062. https://doi.org/10.1093/bioinformatics/bty1054

Smith, S. M., Jenkinson, M., Johansen-Berg, H., Rueckert, D., Nichols, T. E., Mackay, C. E., Watkins, K. E., Ciccarelli, O., Cader, M. Z., Matthews, P. M., & Behrens, T. E. J. (2006). Tract-based spatial statistics: Voxelwise analysis of multi-subject diffusion data. NeuroImage, 31(4), 1487–1505. https://doi.org/10.1016/j.neuroimage.2006.02.024

Styner, M., Oguz, I., Xu, S., Brechbuhler, C., Pantazis, D., Levitt, J. J., Shenton, M. E., & Gerig, G. (2006). Framework for the statistical shape analysis of brain structures using SPHARM-PDM. The Insight Journal, 1071, 242–250.

Tartaglia, M. C., Zhang, Y., Racine, C., Laluz, V., Neuhaus, J., Chao, L., Kramer, J., Rosen, H., Miller, B., & Weiner, M. (2012). Executive dysfunction in frontotemporal dementia is related to abnormalities in frontal white matter tracts. Journal of Neurology, 259(6), 1071–1080. https://doi.org/10.1007/s00415-011-6300-x

Wagner, M., Lorenz, G., Volk, A. E., Brunet, T., Edbauer, D., Berutti, R., Zhao, C., Anderl-Straub, S., Bertram, L., Danek, A., Deschauer, M., Dill, V., Fassbender, K., Fliessbach, K., Götze, K. S., Jahn, H., Kornhuber, J., Landwehrmeyer, B., Lauer, M., … Winkelmann, J. (2021). Clinico-genetic findings in 509 frontotemporal dementia patients. Molecular Psychiatry, 26(10), Article 10. https://doi.org/10.1038/s41380-021-01271-2

Yang, Y., Halliday, G. M., Hodges, J. R., & Tan, R. H. (2017). Von Economo Neuron Density and Thalamus Volumes in Behavioral Deficits in Frontotemporal Dementia Cases with and without a C9ORF72 Repeat Expansion. Journal of Alzheimer’s Disease, 58(3), 701–709. https://doi.org/10.3233/JAD-170002

Zamboni, G., Huey, E. D., Krueger, F., Nichelli, P. F., & Grafman, J. (2008). Apathy and disinhibition in frontotemporal dementia. Neurology, 71(10), 736–742. https://doi.org/10.1212/01.wnl.0000324920.96835.95

Zhang, Y., Schuff, N., Du, A.-T., Rosen, H. J., Kramer, J. H., Gorno-Tempini, M. L., Miller, B. L., & Weiner, M. W. (2009). White matter damage in frontotemporal dementia and Alzheimer’s disease measured by diffusion MRI. Brain, 132(9), 2579–2592. https://doi.org/10.1093/brain/awp071

