## Supplemental tables and figures for "Structural Thalamocortical Network Atrophy in Sporadic Behavioural Variant Frontotemporal Dementia"

### Supplemental Information

Supplemental Table 1. Cortical thickness change between healthy control and bvFTD participants:

| Region | Left |  | Right |  |
| --- | --- | --- | --- | --- |
|  | Beta | p value | Beta | p value |
| bankssts | -0.125 | 0.0236 | -0.066 | 0.1875 |
| caudalanteriorcingulate | -0.204 | 0.0596 | -0.107 | 0.3341 |
| caudalmiddlefrontal | -0.132 | 0.0183 | -0.145 | 0.0075 |
| cuneus | -0.106 | 0.0179 | -0.023 | 0.5958 |
| entorhinal | -0.466 | 0.0062 | -0.395 | 0.0286 |
| fusiform | -0.104 | 0.0276 | -0.034 | 0.4850 |
| inferiorparietal | -0.068 | 0.0920 | -0.030 | 0.4753 |
| inferiortemporal | -0.157 | 0.0049 | -0.068 | 0.2021 |
| isthmuscingulate | -0.162 | 0.0186 | -0.231 | 0.0005 |
| lateraloccipital | 0.041 | 0.4164 | 0.065 | 0.1082 |
| lateralorbitofrontal | **-0.339** | **<0.0001** | **-0.239** | **0.0009** |
| lingual | -0.051 | 0.1882 | -0.018 | 0.6575 |
| medialorbitofrontal | -0.195 | 0.0122 | -0.268 | 0.0022 |
| middletemporal | -0.196 | 0.0022 | **-0.199** | **0.0011** |
| parahippocampal | -0.215 | 0.0993 | -0.177 | 0.0941 |
| paracentral | -0.020 | 0.7269 | -0.063 | 0.1804 |
| parsopercularis | -0.167 | 0.0054 | **-0.166** | **0.0014** |
| parsorbitalis | -0.274 | 0.0041 | **-0.238** | **0.0013** |
| parstriangularis | **-0.255** | **0.0003** | **-0.197** | **0.0011** |
| pericalcarine | -0.041 | 0.2227 | -0.033 | 0.3347 |
| postcentral | 0.004 | 0.9143 | -0.008 | 0.8403 |
| posteriorcingulate | **-0.178** | **0.0002** | -0.159 | 0.0161 |
| precentral | -0.122 | 0.0273 | -0.099 | 0.0159 |
| precuneus | -0.073 | 0.1416 | -0.040 | 0.3030 |
| rostralanteriorcingulate | **-0.334** | **0.0001** | -0.211 | 0.0356 |
| rostralmiddlefrontal | **-0.172** | **0.0061** | -0.181 | 0.0030 |
| superiorfrontal | **-0.184** | **0.0007** | **-0.219** | **0.0003** |
| superiorparietal | 0.006 | 0.8833 | 0.029 | 0.4927 |
| superiortemporal | -0.106 | 0.0941 | -0.137 | 0.0208 |
| supramarginal | -0.127 | 0.0047 | -0.054 | 0.2380 |
| frontalpole | -0.147 | 0.2087 | -0.227 | 0.0567 |
| temporalpole | -0.337 | 0.0149 | -0.208 | 0.0705 |
| transversetemporal | -0.051 | 0.5420 | 0.029 | 0.7396 |
| insula | **-0.278** | **<0.0001** | -0.135 | 0.0129 |

Highlighted regions are significant at a Bonferroni-like corrected threshold of 0.0015 (0.05/34).

Supplemental Table 2. Cortical thickness correlations with clinical and behavioural measures.

| Region | FTLD |  |  |  | FBI all |  |  |  |
| --- | --- | --- | --- | --- | --- | --- | --- | --- |
|  | Right |  | Left |  | Right |  | Left |  |
|  | Beta | p | Beta | p | Beta | p | Beta | p |
| bankssts | -0.010 | 0.280 | -0.004 | 0.394 | -0.004 | 0.382 | -0.004 | 0.394 |
| caudalanteriorcingulate | -0.023 | 0.404 | -0.013 | 0.259 | -0.007 | 0.591 | -0.013 | 0.259 |
| caudalmiddlefrontal | -0.022 | 0.053 | -0.002 | 0.791 | -0.012 | 0.036 | -0.002 | 0.791 |
| cuneus | 0.001 | 0.957 | 0.001 | 0.747 | 0.005 | 0.253 | 0.001 | 0.747 |
| entorhinal | -0.019 | 0.627 | -0.028 | 0.099 | 0.000 | 0.989 | -0.028 | 0.099 |
| fusiform | -0.009 | 0.415 | 0.001 | 0.842 | -0.003 | 0.597 | 0.001 | 0.842 |
| inferiorparietal | 0.005 | 0.559 | -0.005 | 0.271 | 0.004 | 0.306 | -0.005 | 0.271 |
| inferiortemporal | -0.002 | 0.855 | -0.002 | 0.689 | 0.002 | 0.700 | -0.002 | 0.689 |
| isthmuscingulate | -0.016 | 0.136 | -0.003 | 0.624 | -0.003 | 0.600 | -0.003 | 0.624 |
| lateraloccipital | 0.012 | 0.141 | 0.005 | 0.375 | 0.006 | 0.133 | 0.005 | 0.375 |
| lateralorbitofrontal | -0.037 | 0.009 | -0.007 | 0.399 | -0.004 | 0.547 | -0.007 | 0.399 |
| lingual | 0.000 | 0.985 | 0.002 | 0.716 | 0.000 | 0.926 | 0.002 | 0.716 |
| medialorbitofrontal | -0.032 | 0.066 | 0.001 | 0.951 | 0.002 | 0.860 | 0.001 | 0.951 |
| middletemporal | 0.005 | 0.724 | -0.002 | 0.805 | 0.005 | 0.500 | -0.002 | 0.805 |
| parahippocampal | -0.024 | 0.313 | -0.020 | 0.114 | -0.015 | 0.179 | -0.020 | 0.114 |
| paracentral | -0.016 | 0.074 | -0.006 | 0.343 | -0.006 | 0.205 | -0.006 | 0.343 |
| parsopercularis | -0.021 | 0.045 | 0.002 | 0.805 | -0.002 | 0.725 | 0.002 | 0.805 |
| parsorbitalis | -0.037 | 0.004 | -0.007 | 0.481 | -0.006 | 0.334 | -0.007 | 0.481 |
| parstriangularis | -0.021 | 0.045 | -0.005 | 0.472 | -0.005 | 0.318 | -0.005 | 0.472 |
| pericalcarine | -0.007 | 0.253 | 0.000 | 0.935 | -0.001 | 0.841 | 0.000 | 0.935 |
| postcentral | 0.000 | 0.975 | 0.000 | 0.919 | 0.002 | 0.685 | 0.000 | 0.919 |
| posteriorcingulate | -0.031 | 0.020 | -0.003 | 0.466 | -0.009 | 0.163 | -0.003 | 0.466 |
| precentral | -0.014 | 0.078 | -0.011 | 0.061 | -0.008 | 0.030 | -0.011 | 0.061 |
| precuneus | -0.007 | 0.451 | -0.004 | 0.454 | -0.002 | 0.717 | -0.004 | 0.454 |
| rostralanteriorcingulate | -0.033 | 0.168 | -0.007 | 0.278 | -0.001 | 0.953 | -0.007 | 0.278 |
| rostralmiddlefrontal | -0.010 | 0.391 | 0.000 | 0.964 | 0.002 | 0.762 | 0.000 | 0.964 |
| superiorfrontal | -0.029 | 0.024 | -0.006 | 0.243 | -0.009 | 0.112 | -0.006 | 0.243 |
| superiorparietal | -0.001 | 0.919 | -0.001 | 0.782 | 0.001 | 0.907 | -0.001 | 0.782 |
| superiortemporal | -0.005 | 0.712 | -0.005 | 0.542 | -0.001 | 0.851 | -0.005 | 0.542 |
| supramarginal | -0.006 | 0.523 | -0.004 | 0.329 | -0.002 | 0.659 | -0.004 | 0.329 |
| frontalpole | -0.003 | 0.890 | 0.008 | 0.508 | 0.011 | 0.250 | 0.008 | 0.508 |
| temporalpole | -0.004 | 0.868 | 0.009 | 0.563 | -0.001 | 0.971 | 0.009 | 0.563 |
| transversetemporal | -0.007 | 0.596 | -0.006 | 0.429 | -0.008 | 0.211 | -0.006 | 0.429 |
| insula | -0.001 | 0.962 | -0.004 | 0.603 | 0.002 | 0.769 | -0.004 | 0.603 |

Supplemental Table 2. Cortical thickness correlations with clinical and behavioural measures (cont).

|  | FBI 1-10 |  |  |  | FBI 12-20 |  |  |  |
| --- | --- | --- | --- | --- | --- | --- | --- | --- |
|  | Right |  | Left |  | Right |  | Left |  |
| Region | Beta | p | Beta | p | Beta | p | Beta | p |
| bankssts | -0.004 | 0.535 | -0.003 | 0.674 | -0.004 | 0.633 | -0.007 | 0.436 |
| caudalanteriorcingulate | 0.004 | 0.854 | -0.021 | 0.196 | -0.043 | 0.107 | -0.009 | 0.720 |
| caudalmiddlefrontal | -0.009 | 0.264 | -0.003 | 0.732 | -0.023 | 0.031 | 0.000 | 0.977 |
| cuneus | 0.008 | 0.229 | 0.001 | 0.836 | 0.003 | 0.707 | 0.001 | 0.945 |
| entorhinal | -0.005 | 0.857 | -0.044 | 0.058 | -0.005 | 0.902 | -0.030 | 0.389 |
| fusiform | 0.002 | 0.809 | 0.000 | 0.985 | -0.016 | 0.140 | 0.003 | 0.804 |
| inferiorparietal | 0.006 | 0.296 | -0.008 | 0.186 | 0.005 | 0.556 | -0.002 | 0.828 |
| inferiortemporal | 0.002 | 0.732 | -0.001 | 0.883 | 0.000 | 0.966 | -0.009 | 0.419 |
| isthmuscingulate | -0.002 | 0.734 | -0.004 | 0.633 | -0.006 | 0.519 | 0.000 | 0.986 |
| lateraloccipital | 0.011 | 0.050 | 0.008 | 0.310 | 0.000 | 0.958 | 0.003 | 0.771 |
| lateralorbitofrontal | -0.003 | 0.768 | -0.010 | 0.386 | -0.011 | 0.399 | -0.007 | 0.673 |
| lingual | 0.001 | 0.844 | 0.001 | 0.911 | -0.005 | 0.641 | 0.005 | 0.560 |
| medialorbitofrontal | 0.006 | 0.629 | -0.003 | 0.832 | -0.013 | 0.462 | 0.001 | 0.962 |
| middletemporal | 0.006 | 0.492 | -0.003 | 0.737 | 0.003 | 0.815 | -0.003 | 0.790 |
| parahippocampal | -0.021 | 0.184 | -0.039 | 0.024 | -0.026 | 0.249 | -0.010 | 0.694 |
| paracentral | -0.007 | 0.285 | -0.014 | 0.085 | -0.007 | 0.417 | 0.010 | 0.430 |
| parsopercularis | -0.002 | 0.825 | 0.003 | 0.736 | -0.004 | 0.730 | 0.001 | 0.914 |
| parsorbitalis | -0.008 | 0.373 | -0.013 | 0.352 | -0.006 | 0.643 | 0.002 | 0.936 |
| parstriangularis | -0.007 | 0.360 | -0.009 | 0.342 | -0.005 | 0.611 | 0.001 | 0.924 |
| pericalcarine | -0.002 | 0.644 | -0.001 | 0.875 | 0.001 | 0.826 | 0.001 | 0.757 |
| postcentral | 0.002 | 0.733 | 0.003 | 0.664 | 0.001 | 0.881 | -0.005 | 0.603 |
| posteriorcingulate | -0.010 | 0.297 | -0.008 | 0.102 | -0.020 | 0.135 | 0.006 | 0.388 |
| precentral | -0.010 | 0.049 | -0.019 | 0.018 | -0.008 | 0.327 | 0.000 | 0.978 |
| precuneus | -0.002 | 0.749 | -0.007 | 0.295 | -0.001 | 0.878 | 0.002 | 0.837 |
| rostralanteriorcingulate | 0.005 | 0.704 | -0.013 | 0.171 | -0.026 | 0.191 | -0.002 | 0.881 |
| rostralmiddlefrontal | 0.006 | 0.370 | 0.000 | 0.981 | -0.009 | 0.374 | 0.001 | 0.907 |
| superiorfrontal | -0.005 | 0.483 | -0.005 | 0.463 | -0.020 | 0.053 | -0.009 | 0.340 |
| superiorparietal | -0.001 | 0.816 | 0.001 | 0.832 | 0.006 | 0.489 | -0.006 | 0.501 |
| superiortemporal | 0.000 | 0.955 | -0.006 | 0.527 | -0.003 | 0.799 | -0.004 | 0.762 |
| supramarginal | -0.004 | 0.547 | -0.007 | 0.263 | 0.000 | 0.976 | 0.000 | 0.960 |
| frontalpole | 0.009 | 0.522 | 0.003 | 0.862 | 0.019 | 0.325 | 0.021 | 0.377 |
| temporalpole | 0.002 | 0.908 | 0.002 | 0.940 | -0.011 | 0.684 | 0.018 | 0.547 |
| transversetemporal | -0.009 | 0.352 | -0.006 | 0.585 | -0.014 | 0.297 | -0.013 | 0.419 |
| insula | 0.004 | 0.598 | -0.006 | 0.568 | -0.008 | 0.483 | -0.007 | 0.638 |

Supplemental Table 3. DTI correlations with clinical and behavioural measures.

|  | FTLD |  |  | FBI total |  |  | FBI 1-10 |  |  | FBI 12-20 |  |  |
| --- | --- | --- | --- | --- | --- | --- | --- | --- | --- | --- | --- | --- |
|  | Beta | SE | p | Beta | SE | p | Beta | SE | p | Beta | SE | p |
| FA |  |  |  |  |  |  |  |  |  |  |  |  |
| Left ATR | -0.005 | 0.004 | 0.199 | -0.002 | 0.002 | 0.385 | -0.002 | 0.003 | 0.487 | -0.002 | 0.003 | 0.487 |
| Right ATR | **-0.010** | **0.004** | **0.014** | -0.003 | 0.002 | 0.214 | -0.005 | 0.003 | 0.099 | -0.005 | 0.003 | 0.099 |
| Left PTR | -0.002 | 0.003 | 0.446 | -0.002 | 0.001 | 0.246 | **-0.004** | **0.002** | **0.049** | **-0.004** | **0.002** | **0.049** |
| Right PTR | -0.006 | 0.003 | 0.087 | **-0.004** | **0.001** | **0.023** | **-0.005** | **0.002** | **0.026** | **-0.005** | **0.002** | **0.026** |
| MD (× 10^-4^) |  |  |  |  |  |  |  |  |  |  |  |  |
| Left ATR | 0.062 | 0.025 | 0.258 | 0.024 | 0.013 | 0.115 | 0.031 | 0.019 | 0.192 | 0.031 | 0.019 | 0.192 |
| Right ATR | 0.031 | 0.027 | 0.258 | 0.014 | 0.013 | 0.308 | 0.025 | 0.018 | 0.192 | 0.025 | 0.018 | 0.192 |
| Left PTR | 0.034 | 0.019 | 0.088 | 0.017 | 0.010 | 0.115 | 0.025 | 0.014 | 0.079 | 0.025 | 0.014 | 0.079 |
| Right PTR | 0.039 | 0.022 | 0.094 | **0.024** | **0.011** | **0.044** | **0.034** | **0.015** | **0.036** | **0.034** | **0.015** | **0.036** |

#### Validation analyses

SPHARM-PDM (Styner et al., 2006) was used as an alternative shape analysis methodology. Briefly, the segmentations were minimally smoothed with a 1 mm Gaussian kernel and internal holes filled to ensure a spherical topology. Smoothed images were described by spherical harmonics to determine correspondence and then sampled into meshes with 1002 vertices. Surfaces were aligned using a rigid-body Procrustes alignment to a study-specific mean template for each structure. Finally, we calculated the displacement of each vertex for each subject from the mean shape along the vertex normal. These vertex displacement measures were used as the dependent variable in subsequent analyses.

Initially, we utilised SPHARM-PDM vertex analysis as described in the original publication. Here, linear models were analysed at each vertex (vertex displacement ~ ICV + Age + Group) to yield 1002 coefficients and p-values. P-values were subsequently corrected for multiple comparisons using FDR correction. Coefficients are presented in the top left of Supplemental Figure S1. No vertices were significant after FDR correction (not shown).

Subsequently, vertex displacement measures were used in PLS-DA analysis, with control of ICV and age as detailed in the manuscript. Results of coefficients are presented in the top right of Supplemental Figure S1.

Next, we used Deformetrica shape analyses. Here shapes were processed as detailed in the manuscript and two types of analyses were performed. First, like SPHARM-PDM vertex analyses, we created linear models for each momenta direction vector controlling for ICV and age and comparing groups. The coefficient for the group comparison was then projected along the existing momenta directions (“shooting”, in Deformetrica terms) to deform the average model. For visualisation, we observed the displacement along the normal vector between the average and the projected shape, shown in the top right of Supplemental Figure S1. Lastly, we used the momenta as above for PLS-DA classification for the thalami, projected the loadings as described in the manuscript, and visualised the results in the bottom right of Supplemental Figure S1.

The results of various shape analysis and imaging methods are similar in visual comparison.

Supplemental Figure 1.

Group differences using various shape and statistical analyses methods.

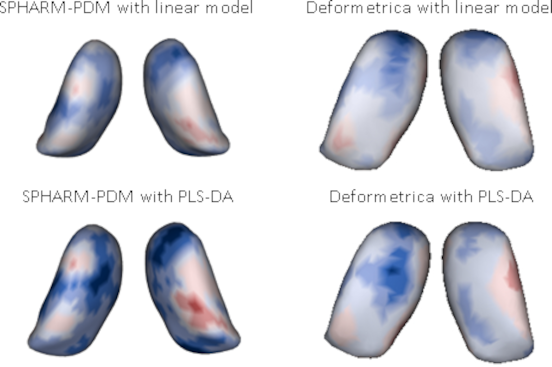

Views of bilateral thalami from superior/top view. Colours indicate displacement relative to an average shape; cooler colours mean inward displacement of the surface, and warmer colours are outward displacement of the surface. Absolute scales are centred around zero with arbitrary magnitude.
